# Correlating hippocampal and amygdala volumes with neuropathological burden in neurodegenerative diseases using 7T postmortem MRI

**DOI:** 10.1101/2024.05.15.24307354

**Authors:** Jr-Jiun Liou, Jinghang Li, Jacob Berardinelli, Hecheng Jin, Tales Santini, Jaehoon Noh, Nadim Farhat, Minjie Wu, Howard Aizenstein, Joseph M Mettenburg, William Yong, Elizabeth Head, Milos Ikonomovic, Tamer Ibrahim, Julia Kofler, the Alzheimer’s Biomarker Consortium on Down syndrome study

## Abstract

Numerous research groups worldwide have focused on postmortem imaging to bridge the resolution gap between clinical neuroimaging and neuropathology data. We developed a standardized protocol for brain embedding, imaging, and processing, facilitating alignment between antemortem MRI, postmortem MRI, and pathology to observe brain atrophy and structural damage progression over time. Using 7T postmortem ex vivo MRI, we explore the potential correlation of amygdala and hippocampal atrophy with neuropathological burden in both Down syndrome (DS) and Alzheimer’s disease (AD) cohorts. Using 7T postmortem ex vivo MRI scans from 66 cases (12 DS and 54 AD) alongside a subset of antemortem scans (n=17), we correlated manually segmented hippocampal and amygdala volumes, adjusted for age, sex, and ApoE4 status, with pathological indicators such as Thal phase, Braak stage, limbic-predominant age-related TDP-43 encephalopathy (LATE) stage, hippocampal sclerosis (HS), and Lewy body (LB) stage. A significant correlation was observed between postmortem and antemortem volumes for the hippocampus, but a similar trend observed for the amygdala did not reach statistical significance. DS individuals exhibited notably smaller hippocampal and amygdala volumes compared to AD subjects. In DS, lower hippocampal and amygdala volumes correlated with more severe Braak stage, without significant associations with Thal phase. LATE and HS pathologies were uncommon in DS cases but trended toward smaller hippocampal volumes. In AD, lower hippocampal volume associated with dementia duration, advanced Thal phase, Braak stage, LATE stage, and HS presence, whereas reduced amygdala volume correlated mainly with severe LATE stage and HS, but not with Thal or Braak stages. No significant LB correlation was detected in either DS or AD cohorts. Hippocampal volume in AD appears influenced by both AD and LATE pathologies, while amygdala volume seems primarily influenced by LATE. In DS, smaller hippocampal volume, relative to AD, appears primarily influenced by tau pathology.

## Introduction

Addressing the resolution gap between 1 mm clinical neuroimaging data and 0.05 mm neuropathology data poses a significant challenge. To bridge this gap, numerous research groups worldwide have focused on postmortem imaging. Over the past years, our team has developed a standardized protocol for brain embedding, imaging, and processing. Additionally, we have designed a 3D-printed brain container with tailored cutting guides for postmortem imaging [1–3]. Utilizing both first [4–6] and second [7, 8] generation of Tic-Tac-Toe head coil radiofrequency system for magnetic resonance imaging (MRI) at 7 Tesla (7T), we have successfully scanned over a hundred brains, generating a substantial volume of high-resolution postmortem imaging data within the University of Pittsburgh.

Postmortem MRI can be conducted in situ or ex vivo. While in situ imaging allows automated image processing, logistical challenges make it impractical. Ex vivo imaging has been approached in various ways, with some studies dissecting the brain into parts [9] or attempting extremely long acquisition times [10]. However, these approaches are impractical for larger cohorts. Researchers who conducted brain imaging directly without any embedding medium [11] or in fluids [9, 10, 12–15] may encounter challenges related to immobilizing the tissue during imaging and aligning the images. Research has shown that agar gel, in contrast to formalin and phosphate-buffered saline, offers superior signal-to-noise and contrast-to-noise ratios as an embedding medium, while also providing stability to the brain [16, 17].

Limbic-predominant age-related TDP-43 encephalopathy (LATE) pathology is a common concurrent neurodegenerative pathology in AD brains, typically initiating in the amygdala before spreading to the hippocampus and neocortex. In radiology-to-pathology correlation studies, LATE pathology at the time of autopsy was associated with smaller amygdala [18] and hippocampal [19] volumes at the time of prior antemortem MRI. Whether similar associations exist in DS, the most common genetic cause of AD, remains unexplored. Investigating the underlying pathologies contributing to hippocampal and amygdala atrophy in DS, especially in a considerably younger population, could offer insights into early AD-related changes in this demographic.

Our study aims to explore the potential correlation of amygdala and hippocampal atrophy with neuropathological burden in both DS and AD using 7T postmortem MRI. A comparative analysis of DS and AD cohorts will help determine whether these observations are associated with AD pathology, LATE pathology, Lewy body (LB) pathology, or a combination thereof.

## Materials and methods

### Participants

All brain donors were either participants in the University of Pittsburgh Alzheimer’s Disease Research Center (ADRC), the Ginkgo Evaluation of Memory Study (GEMS) long-term follow-up cohort, local brain donors to the Pitt Neurodegenerative brain bank or part of the Alzheimer Biomarkers Consortium — Down syndrome (ABC-DS) [20].

The research protocol has been approved by the Committee for Oversight of Research and Clinical Training Involving Decedents at the University of Pittsburgh. It encompasses postmortem brains obtained from December 2019 to May 2023. **Table 1** outlines the fundamental demographics, including age, sex, and race as well as ApoE4 carriership and last clinical diagnosis. The cohort comprises 12 individuals with DS and 54 with AD and related dementias, subsequently labeled as the AD group. Within the AD group, the breakdown includes 18 cases of AD only, 32 cases of AD + Dementia with Lewy bodies (DLB), 1 case of AD + Parkinson’s disease with dementia (PDD), 1 case of DLB, and 2 cases of primary age-related tauopathy (PART). Four individuals in this group were also given an additional diagnosis of argyrophilic grain disease. For DS cases, the most recent DS mental status examination (DSMSE) is available for 10 of the 12 individuals, with the examination conducted 2.0 ± 1.1 years before autopsy. In the Alzheimer’s disease group, 44 of the 54 individuals had their most recent Mini-Mental state examination (MMSE) scores recorded 3.0 ± 2.6 years before death.

**Table 1.** Postmortem brain sample characteristics.

|  | Down syndrome (n=12) | Alzheimer's disease (n=54) |
| --- | --- | --- |
| **Demographic data** |  |  |
| Age (mean ± std) | 60.5 ± 5.0 | 79.7 ± 12.1 |
| Sex female | 41.7% | 48.1% |
| Race |  |  |
| White | 100% | 96.3% |
| Unknown | - | 3.7% |
| **Clinical data** |  |  |
| Last MMSE (mean ± std) | 28.6 ± 21.0 (DSMSE) | 16.5 ± 7.5 |
| MMSE-death interval, y (mean ± std) | 2.0 ± 1.1 | 3.0 ± 2.6 |
| Dementia duration, y (mean ± std) | n/a | 11.1 ± 4.3 |
| ApoE4 status |  |  |
| Carriers | 8.3% | 38.9% |
| Non-carriers | 66.7% | 29.6% |
| Unknown | 25.0% | 31.5% |
| Last clinical diagnosis |  |  |
| No cognitive impairment | 16.7% | 5.6% |
| Mild cognitive impairment | - | 9.3% |
| Alzheimer's disease | 66.7% | 66.7% |
| Other dementia | - | 18.5% |
| Unknown | 16.7% | - |
| **Pathology data** |  |  |
| Postmortem interval, h (mean ± std) | 10.5 ± 8.7 | 8.7 ± 5.5 |
| Fresh brain weight, g (mean ± std) | 1073.7 ± 252.0 | 1164.0 ± 166.9 |
| Thal phase |  |  |
| 0 | - | 3.7% |
| 1 | - | 7.4% |
| 2 | 25.0% | 1.9% |
| 3 | 16.7% | 16.7% |
| 4 | 16.7% | 9.3% |
| 5 | 41.7% | 61.1% |
| Braak NFT stage |  |  |
| 0 | - | 1.9% |
| 1 | - | 3.7% |
| 2 | - | 3.7% |
| 3 | - | 13.0% |
| 4 | 16.7% | 13.0% |
| 5 | 16.7% | 22.2% |
| 6 | 66.7% | 42.6% |
| LATE stage |  |  |
| 0 | 83.3% | 57.4% |
| 1 | - | 11.1% |
| 2 | 16.7% | 31.5% |
| Hippocampal sclerosis | 16.7% | 16.7% |
| Lewy body stage |  |  |
| 0 | 66.7% | 35.2% |
| 1 | - | 13.0% |
| 2 | 16.7% | 18.5% |
| 3 | - | 14.8% |
| 4 | 16.7% | 18.5% |

### Postmortem brain preparation

For postmortem imaging, the fixed brain is embedded into a re-usable 3D printed container [3, 21] that fully fits one adult supratentorial hemisphere without cerebellum or brainstem. The container used in this study consists of four components: a screw cap, domed lid, a cutting guide for coronal slabs, and a container base. Utilizing a Fortus 450 3D printer (Stratasys, USA), we produce these containers using high-density filament polycarbonate (Stratasys, USA). The cutting guides, spaced 0.5 mm apart, facilitate precise coronal brain cutting by neuropathologists.

The average postmortem interval for this cohort is 10.5 hours in individuals with DS and 8.7 hours in those with AD **(Table 1)**. Brain weight measurements were obtained fresh at the time of autopsy prior to dissection. The brainstem was separated at the midbrain level, and the forebrain hemispheres were bisected. The left hemisphere was then fixed in 10% formalin (AD cases) or 4% paraformaldehyde (DS cases) for a minimum of three weeks. To prepare the left hemispheres for imaging, the leptomeninges were removed from the cortical surface to minimize air bubble entrapment. We employed a mixture of 1.5% (w/v) agar (Millipore Sigma, A5431) and 30% sucrose (Fisher, S5-3) to embed the brain in our 3D-printed container. While submerging the brain in the container, we gently massaged it to release any remaining trapped air bubbles. Once the agar began to solidify, the container was sealed with a lid and placed inside the scanner’s head coil for imaging.

### Neuroimaging

Postmortem scans were performed using a 7T human MRI scanner (Siemens Magnetom, Germany) with both first [4–6] and second [7, 8] generation of Tic-Tac-Toe head coil radiofrequency system. Structural imaging included acquisition of both T1-weighted (T1w) MP2RAGE images at a resolution of 0.37 mm and T2-weighted (T2w) SPACE images at a resolution of 0.41 mm, with detailed sequence parameters provided in Table 2. Ex vivo MP2RAGE and SPACE images were registered by rigid registration, and manual segmentation of the hippocampus and the amygdala was performed using ITK-SNAP [22]. To ensure accuracy, all segmentations were evaluated by two authors (J.J.L. and J.N.) before exporting the volumes.

**Table 2.** Postmortem MRI sequence parameters at 7T.

|  | T1w MP2RAGE | T2w SPACE | T2* GRE |
| --- | --- | --- | --- |
| Resolution (mm) | 0.37 | 0.41 | 0.37 |
| Echo time (ms) | 3.57 | 368 | 8.16 |
| Inversion time (ms) | 514 & 2020 | N/A | N/A |
| Repetition time (ms) | 6000 | 3400 | 40 |
| Acceleration factor | 2 | 1 | 2 |
| Number of averages | 2 | 2 | 1 |
| Acquisition time | 32:15 min | 46:36 min | 36:20 min |

Antemortem MR scans were available for 1 DS and 16 AD cases and were retrieved from various studies and clinical radiology records. These scans utilized either a 3T Siemens scanner (Siemens, Germany) or a 3T GE scanner (GE Healthcare, USA). T1-weighted MPRAGE images were acquired with isotropic resolutions ranging from 0.8 to 1.2 mm and acquisition times ranging between 4 and 9 minutes. The interval between postmortem ex vivo and antemortem scans ranged from <1 to 11 years, with an average of 3.8 ± 3.5 years. For antemortem MPRAGE images, hippocampus and amygdala segmentation was performed using FreeSurfer (version 7.3.1, http://surfer.nmr.mgh.harvard.edu/). The extracted antemortem hippocampal and amygdala volumes were correlated with the ex vivo volumes, accounting for discrepancies in absolute values due to the scan-death interval and tissue shrinkage during fixation.

### Neuropathology

Tissue sampling and staining included all brain regions recommended by the 2012 National Institutes of Aging – Alzheimer’s Association (NIA-AA) consensus criteria for the neuropathological evaluation of Alzheimer’s disease [23, 24]. Immunohistochemical staining for beta-amyloid was performed to generate Thal phases [25]. Phospho-Tau staining was performed to determine Braak stage [26]. Modified Bielschowsky stains were used to assess neuritic plaque density by Consortium to Establish a Registry for Alzheimer’s disease (CERAD) criteria [27] for the AD cohort. In the DS cohort, neuritic plaque scores were based on pTau stains. All cases were assigned ABC scores following the NIA-AA criteria, with details of antibodies provided in **Table 3**.

**Table 3.**
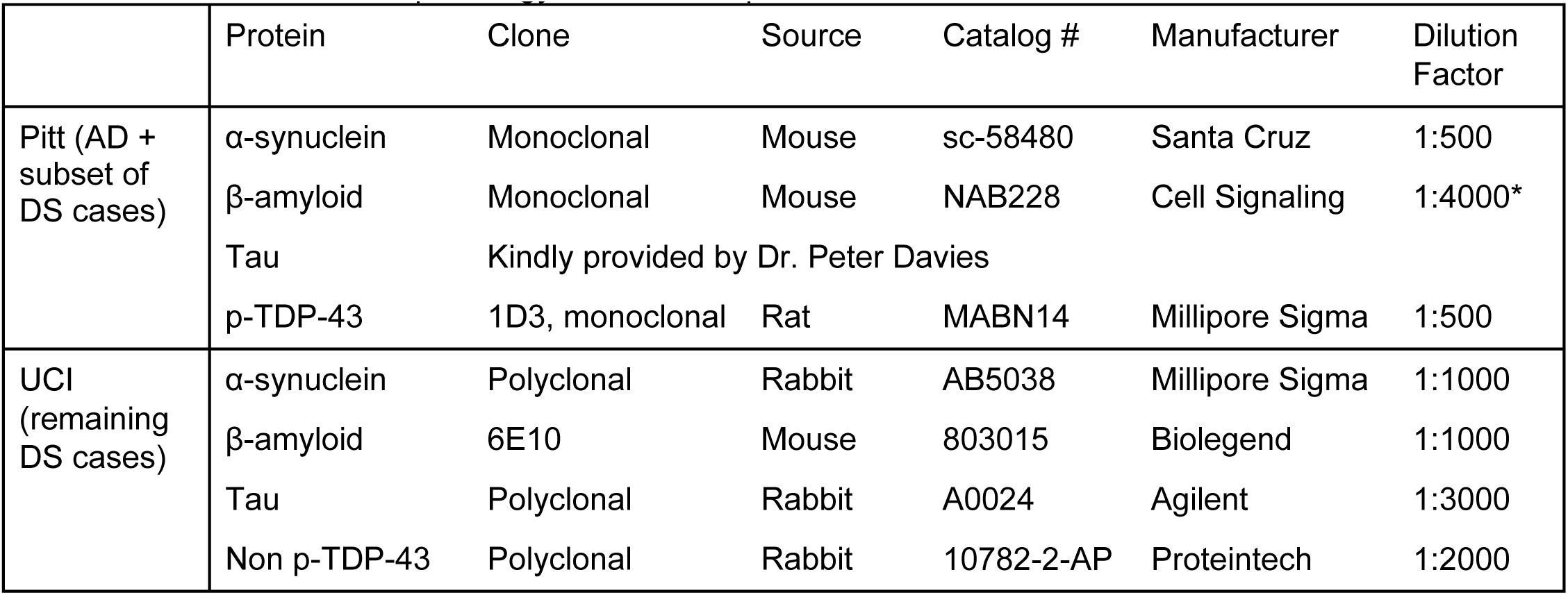
Antibodies for neuropathology. *formic acid pretreatment

TDP-43 immunohistochemistry was carried out on sections from the amygdala, hippocampus-mesial temporal cortex, and midfrontal neocortical regions. TDP-43 positive cases were given a diagnosis of Limbic-Predominant, Age-Related TDP-43 Encephalopathy (LATE). Severity of LATE pathology was assessed by two different methodologies. First, LATE stage was determined following published guidelines by classifying cases based on brain region involvement into stage 1 (amygdala only), stage 2 (stage 1 + hippocampus and/or entorhinal/transentorhinal cortex) and stage 3 (stage 2 + midfrontal cortex) [28]. In addition, the severity of TDP-43 pathology in the following 5 regions was assessed on a semiquantitative scale (none=0, mild=1, moderate=2, severe=3): amygdala, CA1, dentate gyrus, entorhinal/transentorhinal cortex and midfrontal cortex. Stage 2 regions (CA1, dentate gyrus and mesial temporal cortex) were averaged before combining with amygdala and midfrontal cortex score for a final severity score (ranging from 0 to 9). Given the low frequency of TDP43 pathology in the DS cases, severity scores were only obtained for the AD cohort.

Hippocampal sclerosis (HS) evaluation involved assessing two coronal sections, one from the anterior hippocampus and the other from the mid-hippocampus at the level of the lateral geniculate body. Presence of hippocampal sclerosis was determined based on severe neuronal loss and gliosis in CA1 and/or subiculum, disproportionate to AD pathology in the same regions, while blinded to TDP pathology status [23, 29].

Lewy body (LB) stage included stage 0 (negative), stage 1 (olfactory bulb only and brainstem-predominant), stage 2 (amygdala-predominant), stage 3 (limbic), and stage 4 (diffuse neocortical), according to published studies [30–32] and the fourth consensus report of the dementia with Lewy bodies consortium [33] **(Table 1)**.

### Statistical analysis

All statistical analyses were conducted using Prism (version 10.2.0.392) unless specified otherwise. Hippocampal and amygdala volumes were normalized to fresh brain weight and adjusted for age, sex, and ApoE4 carriership using MATLAB (version R2023a). Pearson’s correlation was employed to assess relationships between volume and neuropathological burden with significance set at p < 0.05. Unpaired t-tests were utilized to compare two different groups.

Stepwise regression was conducted using SPSS (version 29.0.2.0) to explore which independent variables contribute the most to postmortem hippocampal and amygdala volumes. The hippocampal or amygdala volume (mm^3^) were the dependent variable; the independent variables included age, sex, ApoE4 carrier status, postmortem interval, fresh brain weight, C score, Thal phase, Braak NFT stage, LATE stage, and LB stage. The adjusted R square, unstandardized coefficients, and associated p-values are reported.

## Results

### Postmortem-antemortem volume correlation

The T1w and T2w images of each brain were registered and used for hippocampal and amygdala segmentation. Segmentations were created manually and quality-inspected **(Figure 1A)**. Among all brains, 17 had antemortem scans with an average postmortem-antemortem scan interval of 3.8 ± 3.5 years. Pearson correlation analysis revealed a significant positive correlation between the postmortem hippocampal volume and that of the antemortem scans (Pearson r = 0.6307, p = 0.0066) **(Figure 1B)**. While a similar trend was observed for the amygdala, it did not reach significance (Pearson r = 0.4210, p = 0.1182) **(Figure 1C)**. It is worth noting that there was one LATE case with a long scan interval of 11 years and a dramatic decrease of amygdala volume between antemortem and postmortem scans (from 2072 to 638 mm^3^) This outlier likely explains the worse correlation for amygdala compared to hippocampal volumes. Overall, these findings indicate that despite the absence of intracranial volume or whole brain volume, the postmortem hippocampal volume derived from manual segmentation accurately represents the antemortem hippocampal volume.

**Figure 1.**
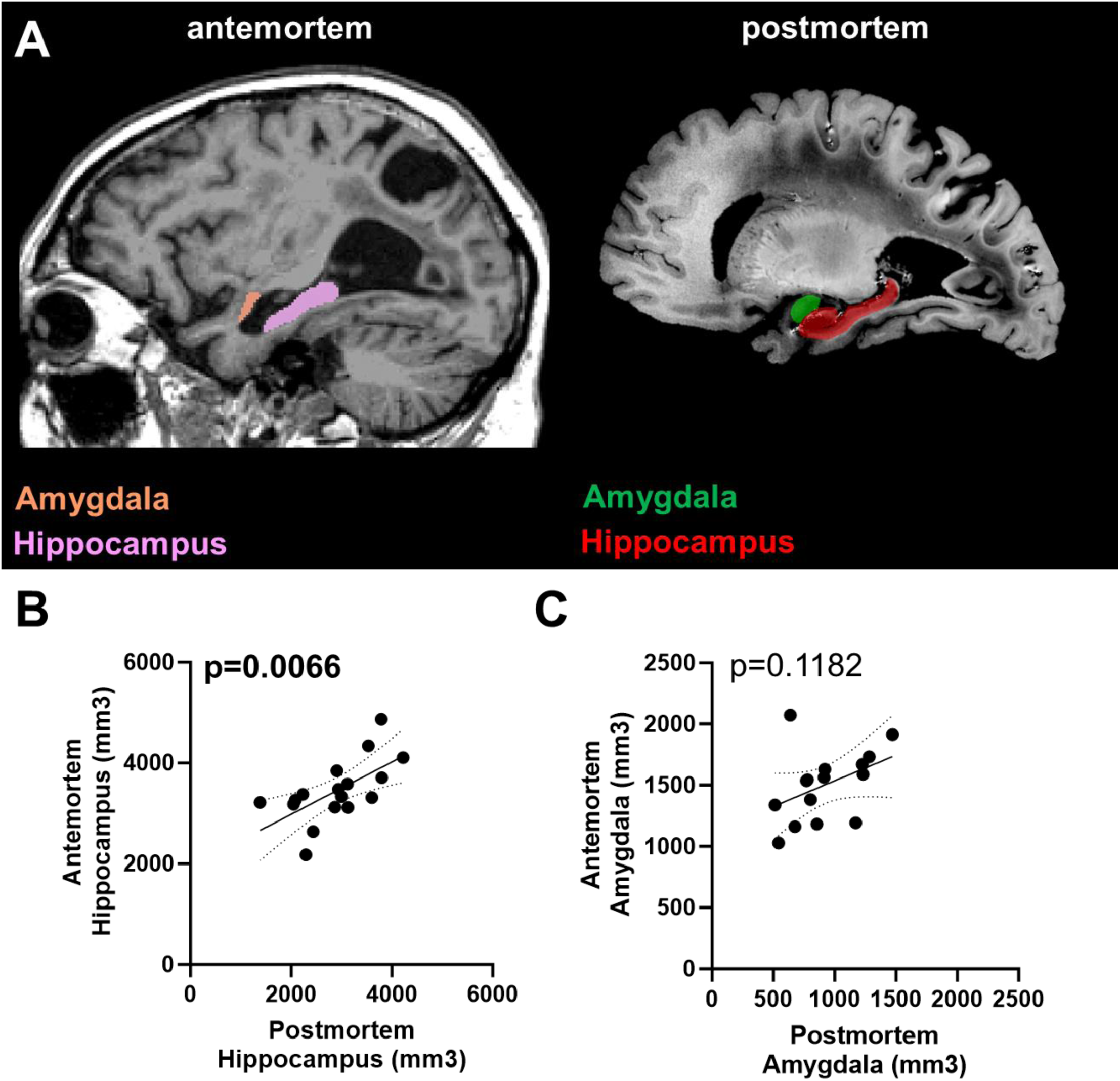
Postmortem-antemortem volume correlation. (A) Postmortem hippocampal and amygdala volumes, obtained from manual segmentations, are compared to a subset of cases with antemortem volumes acquired from FreeSurfer. (B) Significant correlation is observed between postmortem and antemortem hippocampal volumes. (C) However, the correlation between postmortem and antemortem amygdala volumes does not reach significance.

### DS-AD volume comparison

The average age of individuals with DS was approximately 20 years younger than those with AD, with more males and exclusively white participants **(Table 1)**. Regarding ApoE4 carrier status, the majority of DS participants were non-carriers (69.2%), whereas almost an equal number of carriers (38.9%) and non-carriers (29.6%) were reported in the AD cohort. We were unable to obtain postmortem amygdala volumes in four AD cases due to poor delineation in the earliest scans when the protocol was still under development, resulting in an AD sample size of 50 for all amygdala volume analyses. Increased attention to removing entrapped intraventricular air bubbles in the temporal horn improved amygdala delineation in subsequent scans. When comparing hippocampal and amygdala volumes, individuals with DS had significantly lower hippocampal (p=0.0004, **Figure 2A**) and amygdala volumes (p=0.0002; **Figure 2B)** compared to AD cases, after adjusting for age, sex, and ApoE4 carriership.

**Figure 2.**
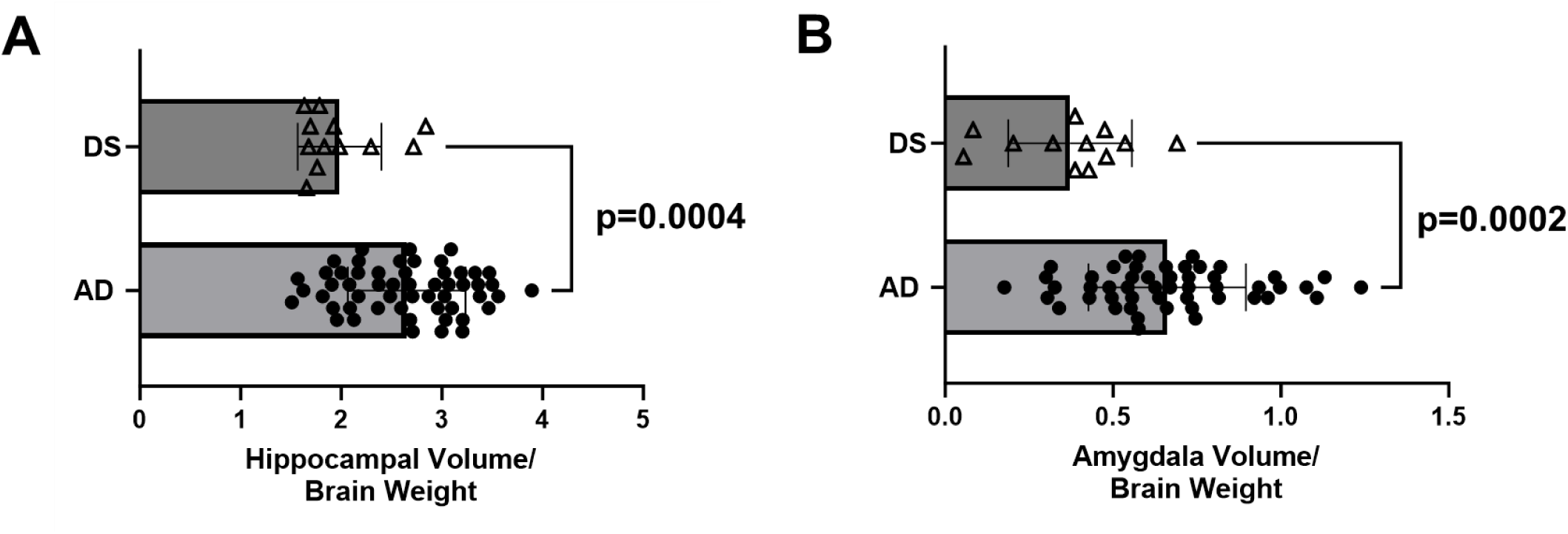
DS and AD volume comparison. (A) Individuals with DS display a significantly lower hippocampal volume than those with AD. (B) Similarly, the amygdala volume is significantly lower in the DS group. All volumes have been normalized to fresh brain weight and adjusted for age, sex, and ApoE4 carriership.

### DS and AD volume-clinical correlation

The most recent DSMSE was obtained on average 2 years prior to autopsy and most DS participants had already developed dementia (61.5%) based on the last clinical diagnosis; however, we had insufficient data to calculate dementia duration for this cohort. In DS, no correlation between DSMSE and hippocampal volume was detected (Pearson r=0.4508, p=0.2233) **(Figure 3A)**, but a trend towards significance was observed in the amygdala (Pearson r=0.6243, p=0.0723) **(Figure 3B)**.

**Figure 3.**
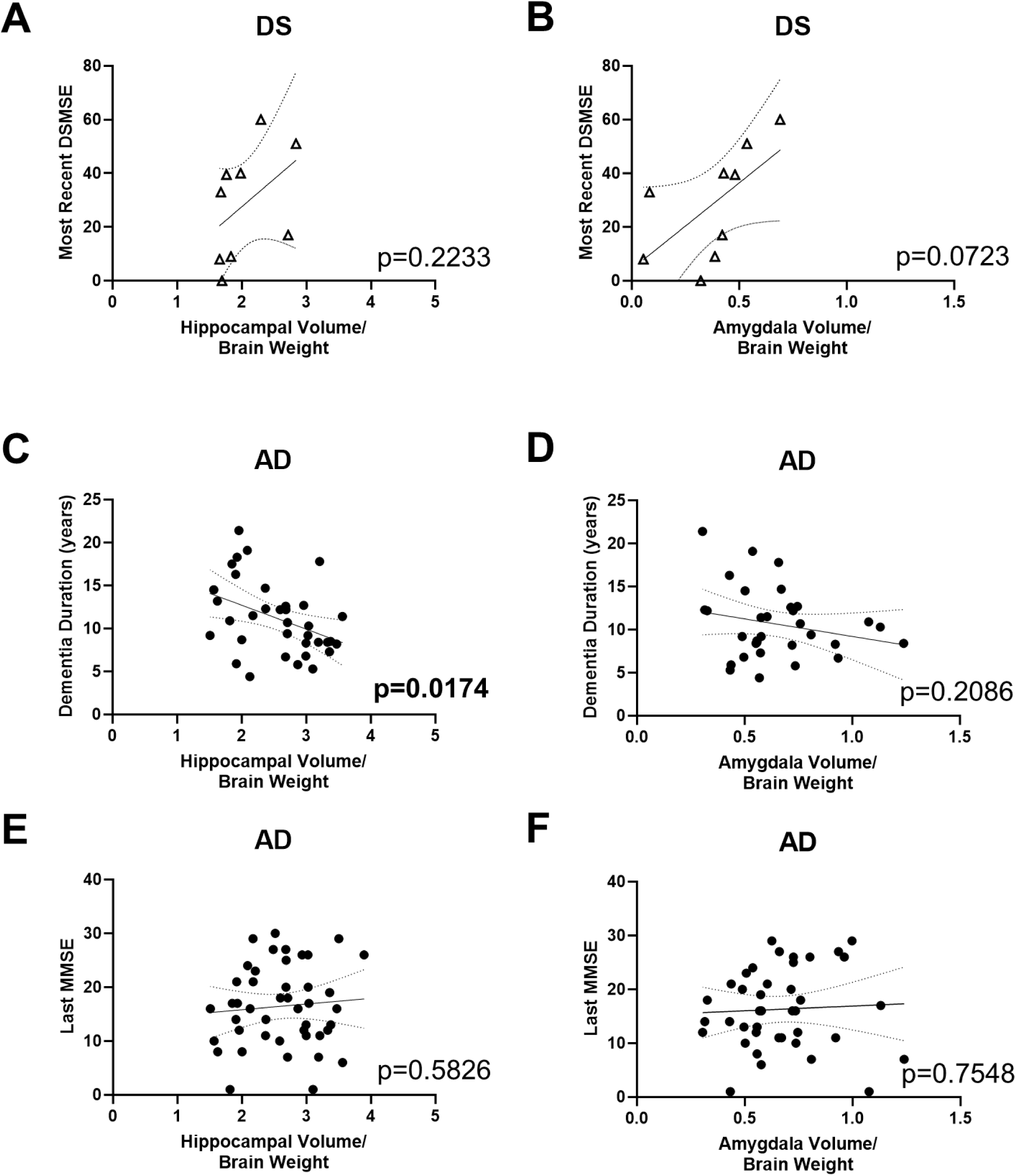
DS and AD volume-clinical correlation. (A) In DS, while no significant correlation between DSMSE and hippocampal volume is observed, (B) a notable trend towards significance is noted in the amygdala. (C) In AD, hippocampal volume shows an inverse correlation with dementia duration, (D) but no correlation is detected in the amygdala. (E) Last MMSE scores do not exhibit correlation with either hippocampal or (F) amygdala volume in AD. All volumes have been standardized to fresh brain weight and adjusted for age, sex, and ApoE4 carriership.

In the AD cohort, 66.7% were clinically diagnosed with probable AD, 18.5% with other forms of dementia, 9.3% with mild cognitive impairment (MCI), and 5.6% had no cognitive impairment. Mean dementia duration was 11 years. In AD, the hippocampal volume had an inverse correlation with dementia duration (Pearson r=-0.3996, p=0.0174), after adjusting for age, sex, and ApoE4 carriership **(Figure 3C)**. However, this correlation did not reach significance in the amygdala (Pearson r=-0.2284, p=0.2086) **(Figure 3D)**. The last MMSE, obtained 3.0 ± 2.6 years prior to autopsy **(Table 1)**, did not show a significant correlation with hippocampal (Pearson r=0.08516, p=0.5826; **Figure 3E)** or amygdala (Pearson r=0.05097, p=0.7548; **Figure 3F)** volume. There was also no correlation between the last MMSE score and the MMSE-autopsy interval (Pearson r=0.1757, p=0.2597).

### Neuropathological burden in DS

The distribution of Thal phase, Braak neurofibrillary tangle (NFT) stage, Lewy body (LB), Limbic-Predominant Age-Related TDP-43 Encephalopathy (LATE), and hippocampal sclerosis (HS) has been summarized in **Table 1**. In the DS group, no significant correlations were detected between Thal phase and hippocampal or amygdala volume **(Figures 4A and 4B)**, but lower hippocampal volume (Pearson r=-0.7481, p=0.0051) **(Figure 4C)** and amygdala volume (Pearson r=-0.6915, p=0.0127) **(Figure 4D)** correlated with more severe Braak NFT stage. In the two DS cases that exhibited LATE pathology and hippocampal sclerosis, trends toward smaller hippocampal and amygdala volumes were observed but not reaching significance **(Figures 4E, 4F, 4G, and 4H).** In the four DS cases that exhibited LB, two with amygdala-predominant LB and two with diffuse neocortical LB, no significant difference in hippocampal volume (p=0.5262) **(Figure 4I)** or amygdala volume (p=0.9124) **(Figure 4J)** was observed when compared to the non-LB group.

**Figure 4.**
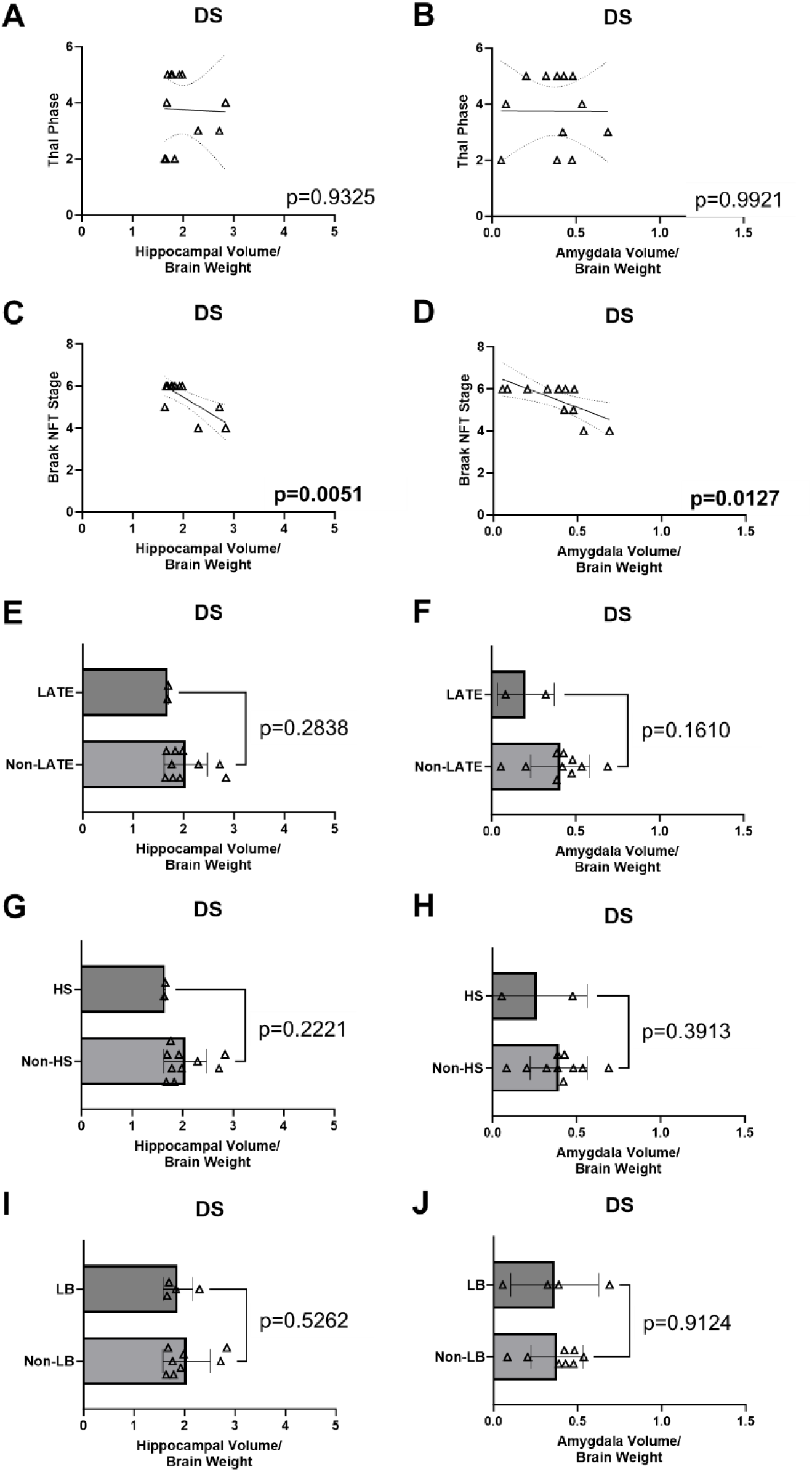
DS volume-neuropathology correlation. (A) No correlation is observed between hippocampal volume and Thal phase. (B) Similarly, no correlation is detected between amygdala volume and Thal phase. (C) Both hippocampal and (D) amygdala volumes correlate with Braak NFT stage. (E) Trends toward smaller hippocampal volume are observed in two DS cases with LATE pathology. (F) However, no significant difference is found in amygdala volume between LATE and non-LATE groups. (G) In two DS cases with HS, trends toward smaller hippocampal volume are noted, but (H) no significant difference is seen in amygdala volume. (I) No distinction is observed between LB and non-LB cases in terms of hippocampal volume, and (J) similarly, no difference is detected in amygdala volume. All volumes have been standardized to fresh brain weight and adjusted for age, sex, and ApoE4 carriership.

### Neuropathological burden in AD

In AD, lower hippocampal volume correlated with more severe Thal phase (Pearson r=-0.4120, p=0.0020), Braak NFT stage (Pearson r=-0.3565, p=0.0081), LATE stage (Pearson r=-0.4170, p=0.0017), and was associated with the presence of HS (p<0.0001) **(Figures 5A, 5C, 5E and 5G)**. Conversely, lower amygdala volume correlated with more severe LATE stage (Pearson r=-0.3390, p=0.0160) and was associated with HS (p=0.0139) but did not correlate with Thal phase (Pearson r=-0.1014, p=0.4833) or Braak stage (Pearson r=0.01762, p=0.9034) **(Figures 5B, 5D, 5F and 5H).** No significant correlation with LB stage was detected in the hippocampus (p=0.7953) or the amygdala (p=0.3684;**(Figure 5I and 5J)**.

**Figure 5.**
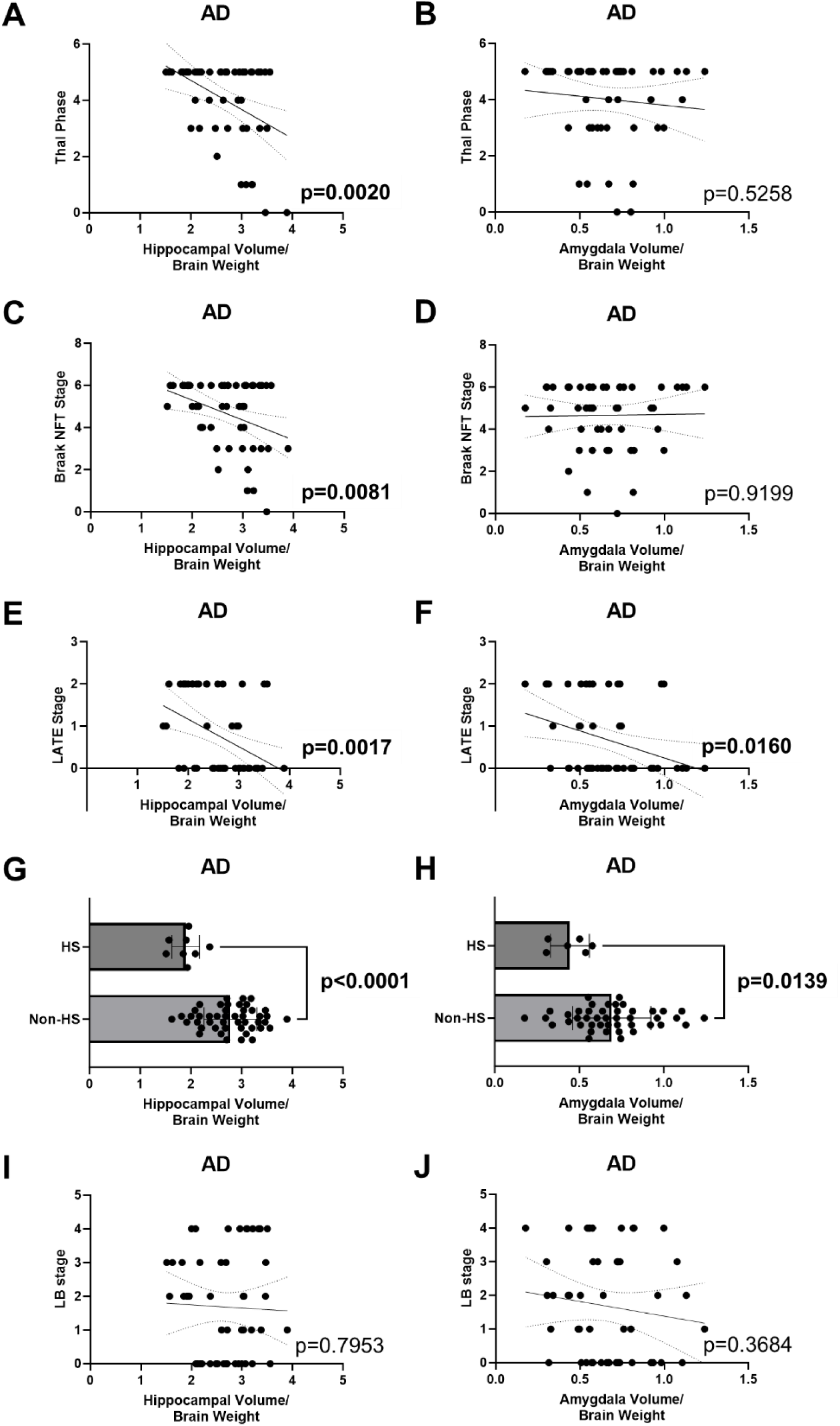
AD volume-neuropathology correlation. (A) The hippocampal volume exhibits correlations with Thal phase, (C) Braak NFT stage, (E) LATE stage, and (G) the presence of HS. (F) Additionally, the amygdala volume correlates with LATE stage and (H) the presence of HS. However, no significant correlations are detected between (B) Thal phase or (D) Braak NFT stage, or (J) LB pathology with the amygdala volume. (I) The hippocampal volume in the LB limbic subtype is significantly lower than in LB neocortical diffuse cases. All volumes have been standardized to fresh brain weight and adjusted for age, sex, and ApoE4 carriership.

We conducted stepwise regression analyses to determine the independent variables contributing to the variance in hippocampal and amygdala volumes, which were normalized to fresh brain weight and adjusted for age, sex, and ApoE4 status in AD. The stepwise regression revealed that C score and LATE stage are the predictors **(Figure 6A)**. The C score accounted for 17.6% of the variance, followed by the LATE stage with an additional 8.1%, totaling 25.7% of the variance in postmortem hippocampal volume. In the case of postmortem amygdala volume (**Figure 6B**), LATE stage emerged as the sole predictor in the stepwise regression model, accounting for 8.7% of the variance in amygdala volume.

**Figure 6.**
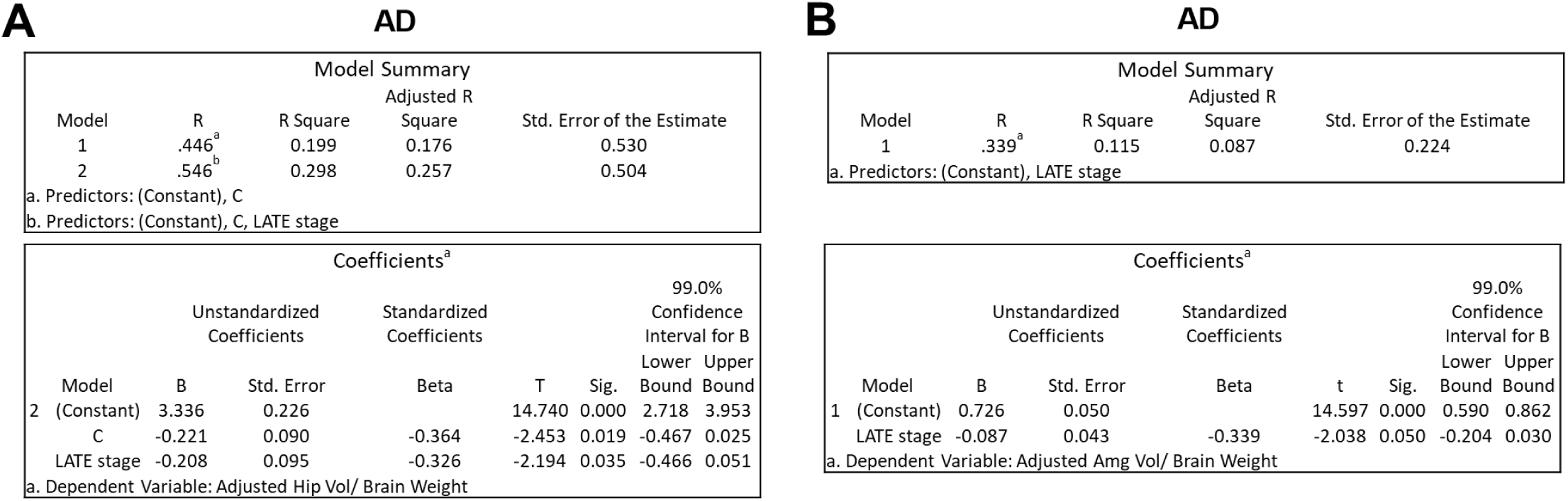
In AD, stepwise regression shows that. (A) both C score and LATE stage are the predictors accounting for 25.7% of the variance in adjusted hippocampal volume; and that (B) LATE stage is the only predictor in adjusted amygdala volume.

## Discussion

We implemented a postmortem ex vivo MRI protocol to investigate the correlation between hippocampal and amygdala volumes and neuropathological burden in neurodegenerative diseases. The significant correlation found between antemortem and postmortem hippocampal volumes suggests the reliability of manually segmented volumes, particularly in cases where intracranial or whole brain volume data is unavailable. While we observed a trend towards correlation between antemortem and postmortem volumes in the amygdala, this correlation did not reach significance, likely due to two primary reasons. Delineation of anatomic structures in the anterior medial temporal lobe was challenging in three early scans, while we were still refining the imaging protocol. Secondly, an outlier case with a scan interval of 11 years and severe amygdala volume loss over the years may have contributed to this lack of significance.

In our analysis, we observed significantly smaller hippocampal and amygdala volumes in individuals with DS compared to those with AD, which is consistent with findings from previous studies [34–39]. Interestingly, research on children with DS revealed no differences in amygdala volumes between DS and control groups, but hippocampal volumes were notably smaller in the DS group [40]. This suggests that decreased hippocampal volumes in DS may also be attributed to early developmental differences in addition to neurodegenerative changes. Hippocampal volumes have been previously shown to correlate with cognitive measures (DSMSE scores) in DS cases [37]. While we observed trends in the same direction for both hippocampal and amygdala volumes, our analyses did not reach significance, likely due to the overall low number of cases in our DS cohort. In contrast, within our AD cohort, hippocampal volume exhibited an inverse correlation with dementia duration, a novel finding in our study and consistent with our previously reported association of hippocampal sclerosis and LATE pathology with duration of cognitive symptoms in a non-overlapping cohort of cases [41]. However, we did not find a significant correlation between hippocampal or amygdala volume and the last MMSE score, even after adjusting for the MMSE-autopsy interval, contrary to findings in some prior studies [42–46], likely due to differences in sample size and group ratios.

Our neuropathological results showed that both hippocampal and amygdala volumes correlated with Braak NFT stage in the DS cohort, after adjusting for age, sex, and ApoE4 carriership. However, no correlations with Thal phase were detected. While LATE pathology was not common in the DS cohort, its frequency was consistent with findings from other studies in younger populations, reporting TPD43 pathology in 9-18% of DS, familial and early-onset sporadic AD cases [47–50]. Despite the low numbers, we observed a trend towards small hippocampal and amygdala volumes in the LATE-positive DS cases. Similar trends were observed for the two DS cases that exhibited HS. Interestingly, both HS cases in this cohort were negative for TDP43 pathology, which is consistent with other studies where half of the DS cases with HS were non-LATE cases [48, 49]. We also identified four DS cases with Lewy body pathology (30.8%), which falls within the published prevalence range of 8-50% [51, 52].

In AD, our neuropathological findings are consistent with previous research, demonstrating an inverse correlation between hippocampal volume and the Thal phase [53, 54], Braak NFT stage [55–57], and LATE stage [43, 58, 59]. Notably, a study suggested that including the TDP-43 stage could explain approximately 3% of the variance in hippocampal volume [59]. Similarly, in our stepwise regression analysis, the LATE-stage accounts for 8.1% of the variance in the adjusted hippocampal volume and 8.7% of the variance in amygdala volume. As expected and consistent with previous findings, we observed lower hippocampal volumes in cases with HS compared to those without, [60] [61, 62]. Interestingly, TDP/HS pathology seems to have a stronger association with hippocampal volume than AD pathology in older individuals with AD [59]. Compared to that study with a mean age at death of around 90 years, the average age in our AD cohort was about a decade lower, which may explain why AD and LATE pathologies had comparable effect sizes in our cohort. Furthermore, we noted an association between amygdala volume and LATE stage. In our stepwise regression analysis, the LATE stage emerged as the sole predictor for postmortem amygdala volume. Regarding the contribution of LB, antemortem imaging studies have reported that less severe hippocampal atrophy is associated with dementia with LBs when compared to AD [63, 64]. We did not observe any significant differences in hippocampal or amygdala volume between LB and non-LB cases, nor did we find an association between volumes and LB stage. These results align with previous findings of comparable amygdala volumes between AD and DLB [46].

This exploratory study is limited by the small sample size, particularly affecting analyses of our DS cohort. Previous studies have indicated that female individuals with DS tend to experience longer durations of dementia [65] and exhibit higher tau burden in vivo [66], while males with DS and the ApoE4 allele display elevated white matter hyperintensity volumes in the occipital lobe [67]. Due to the small sample size of our DS cohort, we had to defer exploration of sex differences in neuropathological burden to follow-up studies when additional cases in the ABC-DS cohort will have come to autopsy. Despite this limitation, our study provides novel insights into differential impact of neurodegenerative pathologies on regional volumes between DS and sporadic AD cases.

## Conclusions

Our 7T postmortem MRI protocol produced good alignment of postmortem volumes with antemortem findings and revealed correlations between volume measures and neuropathological burden. In AD, hippocampal volume is influenced by both AD and LATE pathologies, whereas amygdala volume appears to be influenced primarily by LATE. In DS, hippocampal volume is smaller than in AD and primarily influenced by tau pathology.

## Data Availability

All data produced in the present work are contained in the manuscript

## Abbreviations

3D: three-dimensional
AD: Alzheimer’s disease
ApoE4: apolipoprotein E allele 4
DS: Down syndrome
DSMSE: Down syndrome mental status examination
GRE: GRadient Echo
HS: hippocampal sclerosis
LATE: limbic-predominant age-related
TDP-43: encephalopathy
LB: Lewy body
MMSE: mini mental state examination
MP2RAGE: Magnetization Prepared – RApid Gradient Echo
MRI: magnetic resonance imaging
NFT: neurofibrillary tangle
SPACE: Sampling Perfection with Application optimized Contrast using different flip angle Evolution
TDP-43: TAR DNA-binding protein 43
PMI: postmortem interval

## Acknowledgement

The authors express their sincere gratitude to all patients and their dedicated service providers and families who generously volunteered as participants in these studies. Additionally, the authors extend their heartfelt thanks to the staff members who helped with autopsy coordination and study administration, including Sierra Wright, Joni Vander Bilt, and the autopsy coordination team of the Pitt ADRC. Special appreciation is also extended to the other principal investigators at the University of Pittsburgh for their collaboration and for sharing in vivo scans with the research team. This research was supported in part by the University of Pittsburgh Center for Research Computing, RRID:SCR_022735, through the resources provided. Specifically, this work used the H2P cluster, which is supported by NSF award number OAC-2117681.

#Alzheimer’s Biomarker Consortium-Down syndrome (ABC-DS) Investigators: Beau M. Ances, MD, PhD; Howard F. Andrews, PhD; Karen Bell, MD; Rasmus M. Birn, PhD; Adam M. Brickman, PhD; Peter Bulova, MD; Amrita Cheema, PhD; Kewei Chen, PhD; Bradley T. Christian, PhD; Isabel Clare, PhD; Lorraine Clark, PhD; Ann D. Cohen, PhD; John N. Constantino, MD; Eric W. Doran, MS; Anne Fagan, PhD; Eleanor Feingold, PhD; Tatiana M. Foroud, PhD; Benjamin L. Handen, PhD; Sigan L. Hartley, PhD; Rachel Henson, PhD; Christy Hom, PhD; Lawrence Honig, MD; Sterling C Johnson, PhD; Courtney Jordan, RN; M. Ilyas Kamboh, PhD; David Keator, PhD; William E. Klunk, MD PhD; William Charles Kreisl, MD; Sharon J. Krinsky-McHale, PhD; Florence Lai, MD; Patrick Lao, PhD; Charles Laymon, PhD; Joseph Hyungwoo Lee, PhD; Ira T. Lott, MD; Victoria Lupson, PhD; Mark Mapstone, PhD; Chester A. Mathis, PhD; Davneet Singh Minhas, PhD; Neelesh Nadkarni, MD; Sid O’Bryant, PhD; Deborah Pang, MPH; Melissa Petersen, PhD; Julie C. Price, PhD; Margaret Pulsifer, PhD; Michael Rafii, MD, PhD; Eric Reiman, MD; Batool Rizvi, MS; Herminia Diana Rosas, MD; Marwan N. Sabbagh, MD; Nicole Schupf, PhD; Wayne P. Silverman, PhD; Dana L. Tudorascu, PhD; Rameshwari Tumuluru, MD; Benjamin Tycko, MD, PhD; Badri Varadarajan, PhD; Desiree A. White, PhD; Michael A. Yassa, PhD; Shahid Zaman, MD, PhD; Fan Zhang, PhD.

